# Brief assessment of cognition in immune effector cell-associated neurotoxicity syndrome

**DOI:** 10.1101/2025.06.05.25329077

**Authors:** Baibin (Ben) Wang, Valeriya Kuznetsova, Eunice Lui, Anna Li, Samantha M Loi, Stefanie Roberts, Fiore D’Aprano, Tomas Kalincik, Simon J Harrison, Mark R Dowling, Michael Dickinson, Mark Walterfang, Dennis Velakoulis, Mary Ann Anderson, Charles B Malpas

## Abstract

**Background:** Immune-effector cell-associated neurotoxicity syndrome (ICANS) is a common acute adverse event of chimeric antigen T-cell (CAR–T) therapy associated with a range of neurological and cognitive symptoms. Recent evidence has highlighted that cognitive changes in ICANS may predate changes in the current standard cognitive screening instrument. This study aimed to develop a novel screening instrument sensitive to ICANS-related cognitive dysfunction.

**Methods:** Patients were administered a 21-item bedside cognitive screening instrument (the Neuropsychiatry Unit Cognitive Assessment Tool) followed by an inpatient neuropsychological assessment. A cognitive screening index was computed by summing the beside items that maximally differentiated between ICANS and non-ICANS patients. A separate neuropsychological index was also derived.

**Results:** Among the 50 patients included, 26 (52%) had ICANS at assessment. Six screening tests and five neuropsychological tests demonstrated acceptable clinical utility for indices computation (AUC>0.70). ICANS patients had lower scores on both the cognitive screening index (*p*<.001) and the neuropsychological index (*p*< .001) than non-ICANS patients. The cognitive screening index (AUC=0.80) and the neuropsychological index (AUC=0.81) achieved acceptable clinical utility. Although there was no significant difference between the AUCs of the two indices (*p*=.765), hierarchical logistic regression showed that the neuropsychological index improved the classification of ICANS patients (*p*<.05).

**Conclusion:** The cognitive screening index can reliably discriminate between ICANS and non-ICANS patients, but neuropsychological examination provides more detailed cognitive characterisation in ICANS. Further validation of the cognitive screening index will inform practice in this emerging area of clinical need.

## Introduction

The immune effector cell-associated neurotoxicity syndrome (ICANS) is a common acute adverse event associated with chimeric antigen receptor T-cell (CAR-T) therapy, which is part of standard care for several types of refractory haematological malignancies.^1,2^ A broad range of cognitive, neurological, and psychiatric symptoms have been described in ICANS,^3^ with most symptoms typically occurring within 1**–**3 weeks after an infusion of CD19-directed CAR-T cells in patients treated for B-cell lymphoma or B-cell leukaemia.^4^ Delayed atypical clusters of symptoms, include cranial nerve palsies and movement disorders, have been reported following B-cell maturation antigen directed CAR-T products used for treatment of multiple myeloma. These symptoms have been defined as movement and neurocognitive treatment-emergent adverse events (MNTs) and are thought to be distinct from the more typical ICANS presentation.^5,6^

Detection of ICANS is complicated by the diverse range of symptoms.^3,7^ Kazzi et al^8^ systematically reviewed acute cognitive symptoms following CAR-T therapy and found that the presence and nature of cognitive impairment (e.g. attention deficits, executive dysfunction, language disturbance, and memory deficits) varied across studies. At a study level, the cognitive syndrome of ICANS also varied across patients. For example, Belin et al^9^ reported a sample of 43 ICANS patients with cognitive symptoms ranging from aphasia, cognitive slowness, agraphia, executive syndrome, and memory disorders. Yuen et al^10^ described a cohort of eight ICANS patients with symptoms, such as disorientation, dysgraphia, dysnomia, and inattention.

The detection and grading of ICANS is based on the consensus criteria outlined by the American Society of Transplantation and Cellular Therapy (ASTCT)^3^. The immune effector cell-associated encephalopathy (ICE) is a brief beside-screening test that determines the severity of cognitive symptoms in ICANS in the ASTCT criteria, with lower scores indicating greater severity. Recent studies have questioned the sensitivity of the ICE score in detecting early cognitive symptoms of ICANS. For example, Herr et al^11^ found that multiple patients were unable to follow multi-step commands prior to a reduction in the ICE score. Yuen et al^10^ noted that the ICE does not examine executive function, which they postulated could be disrupted in ICANS. At a content-validity level, the ICE does not capture several crucial domains of cognition, such as executive function, processing speed, or visual-constructional functions.^12^ The lack of items in the aforementioned cognitive domains raises questions regarding the sensitivity of the ICE assessment to detect early subtle cognitive changes.^13^

The primary aim of this study was to derive a new brief cognitive screening instrument sensitive to ICANS-related cognitive change. We also aimed to compare the performance of this new screening instrument to formal neuropsychological tests.

## Methods

### Patients

This was a prospective cohort study of adult patients admitted to a clinical haematology ward at Peter MacCallum Cancer Centre (PMCC) in Melbourne, Australia for routine post-CAR-T monitoring. Patients were eligible if they received commercial CAR-T therapy between June 2021 and March 2024 for treatment of B-cell lymphoma or B-cell leukaemia. Patients were included if they were proficient in English and had no pre-existing significant neurological illness.

### Control participants

Data from 118 healthy control participants were included for use in secondary psychometric analyses. These data were obtained from a larger validation study of the 21-beside screening items and are described elsewhere.^14^ Raw item-level data were provided by the study authors, which we included to investigate the psychometric properties of the derived cognitive screening index, as described below.

### Clinical information

Patients meeting the above criteria were prospectively recruited to the research project prior to CAR-T infusion and inpatient admission. Written informed consent was obtained. Clinical information was obtained from routine inpatient neuropsychological assessments at PMCC. Other clinical information, including demographic characteristics, adverse events, presenting symptoms, findings from routine ICE assessments, grades of cytokine release syndrome (CRS) and ICANS, and details of inpatient medical interventions were obtained from the electronic medical record. Diagnosis of ICANS was made by a multi-disciplinary haematology-led team according to the ASTCT criteria.^3^ The ICE assessment was routinely administered by the nursing staff to monitor for cognitive and neurological changes. The ICE test was scores 0-10, with lower scores indicating greater severity of cognitive symptoms. This study received approval from the PMCC Human Research Ethics Committee (21/145).

### Neuropsychological Evaluation

All patients underwent routine neuropsychological evaluation during their inpatient admission. For patients who developed ICANS, the neuropsychological examination, completed immediately after the date of onset, was included in this study as being the most representative of the cognitive manifestation of early ICANS. For patients who did not develop ICANS, neuropsychological evaluation was performed at approximately day nine post-infusion.

The Neuropsychiatry Unit Cognitive Screening Tool (NUCOG)^14^ was routinely administered to all patients. The NUCOG is a 21-item screening instrument that examines aspects of attention, executive function, memory, language, and visuoconstructional function. The total score of the NUCOG represents general cognitive status and ranges from 0 to 100. A total score of <80 has been shown to detect patients at risk of cognitive impairment.^14^ All NUCOG items were analysed individually to determine which specific items maximally distinguished patients with ICANS from those without.

Formal cognitive tests were administered as part of the routine inpatient neuropsychological evaluation. The specific tests were chosen by the treating neuropsychologists, but generally consisted of the Mental Control subtest from the Weschler Memory Scale-Third Edition (WMS-III),^15^ the oral version of the Symbol Digital Modalities Test (SDMT-Oral),^16^ the Hopkins Verbal Learning Test-Revised (HVLT-R),^17^ the Controlled Oral Word Association Test (COWAT),^18^ the Category Fluency Test (CFT),^19^ the Trail Making Test (TMT),^20^ the copy trial of the Rey-Osterrieth Complex Figure (ROCF),^21^ and the Taylor Complex Figure Test (TCFT).^22^ Neuropsychological test scores were converted to standardised z-scores using demographically-adjusted normative data. Full details of the normative data are included in supplementary materials.

### Statistical analyses

All statistical analyses were performed using R (4.3.1), Jamovi (2.5.3), and JASP (0.18.3) software packages. Care was taken to inspect all data and models for statistical assumptions. Non-parametric methods were used in cases where statistical assumptions were violated. Pair-wise deletion was used in cases of missing data.

Receiver operator characteristic curves (ROC) were computed for all screening test items to determine the ability of the specific item to accurately detect ICANS. The area under the curve (AUC) was computed for each score to summarise classification accuracy. Confidence intervals (CIs) at the 95% level were computed using stratified bootstrapping with 10,000 replicates using the *pROC* package.^23^ Classification accuracy was considered acceptable if the AUC was greater than 0.7 and if the CIs did not capture the null hypothesis value of 0.5.^24,25^ A screening index was computed by summing the scores of acceptable items. An alternative cognitive screening index was then computed by summing the number of abnormal individual items contributing to the cognitive screening index. Given the absence of item level data, the threshold for abnormal scores was determined for each item by computing Youden’s *J* statistic. The same procedure was then repeated for individual neuropsychological tests and a neuropsychological index was derived by computing the mean z-score across acceptable tests. For each patient, an alternative index was computed by summing the number of z-scores < -1.5 SD.

The derived indices were analysed using ROC curves and Youden’s *J* statistic was computed to identify the optimal cut-off for each index. For reliability indices, the greatest lower bound (GLB) coefficient was computed and values > 0.7 were considered acceptable.^26^ The classification performance of all derived indices was compared by testing the difference in AUC using permutation testing.^27^ Hierarchical logistic regression was used to investigate whether the neuropsychological index improved detection of ICANS over and above the cognitive screening index alone. Finally, Welch’s independent samples t-tests were used to investigate mean differences of the derived indices between the ICANS and non-ICANS groups. Spearman’s rank-order correlations with Bonferroni correction were computed to examine the convergence of the derived indices to key clinical measures, including the total length of inpatient stay, last administered corticosteroid dosage by weight (mg/kg), and the ICE score at assessment.

### Data Accessibility Statement

Anonymised data can be shared on reasonable request from qualified investigators.

## Results

As outlined in the supplementary materials (S1), 50 patients were included in the study. Most of the included patients were male (*n* = 31, 62%). The mean age of male patients was 61.45 years (*SD* = 12.93, range = 19-80), and the mean age of female patients was 61 years (*SD* = 11.35, range = 33-75). Nineteen patients developed ICANS with a maximum grade of 1, three patients developed ICANS with a maximum grade of 2, ten patients developed ICANS with a maximum grade of 3, and one patient developed ICANS with a maximum grade of 4. Table 1 outlines the clinicodemographic characteristics of the cohort.

**Table 1.**
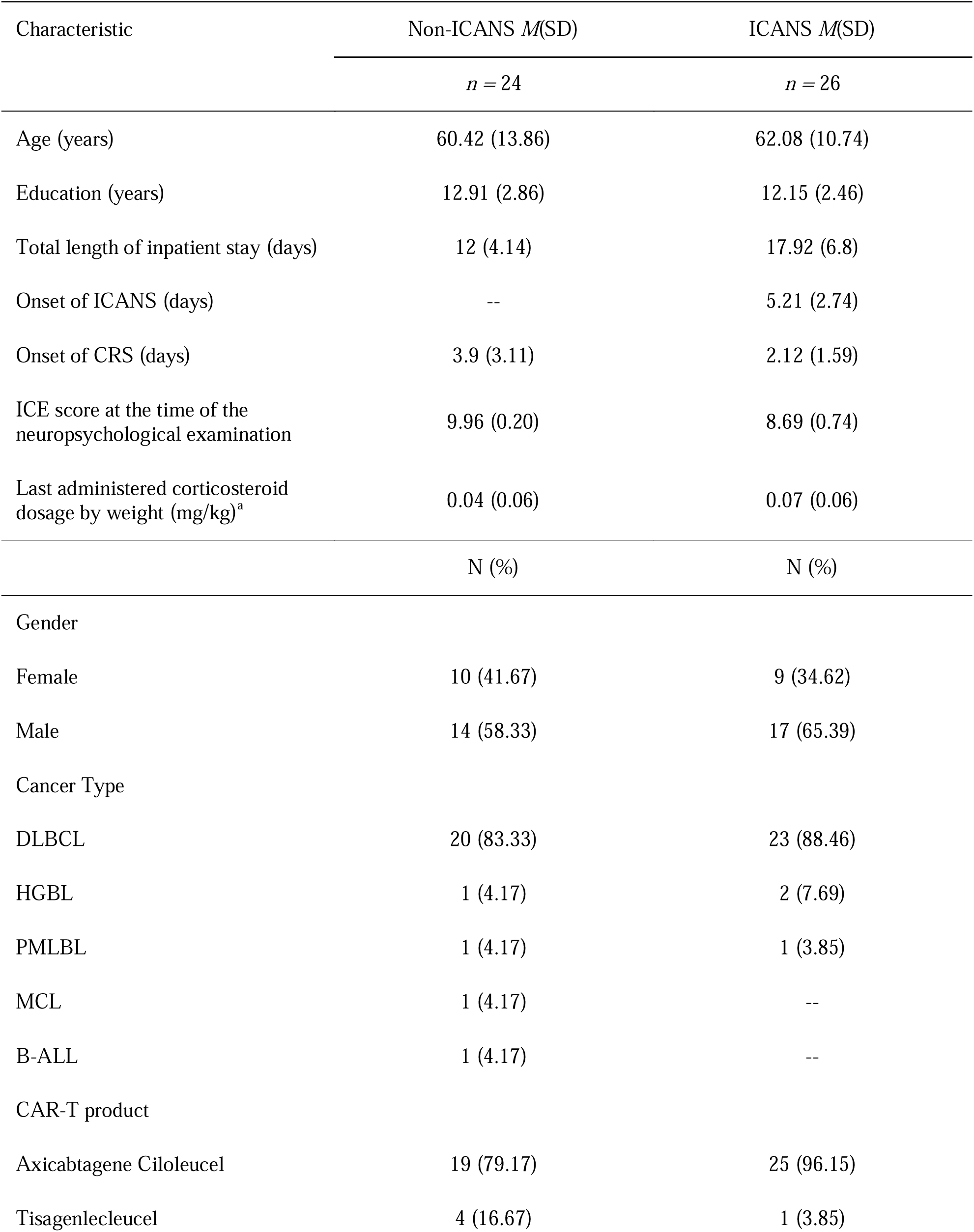
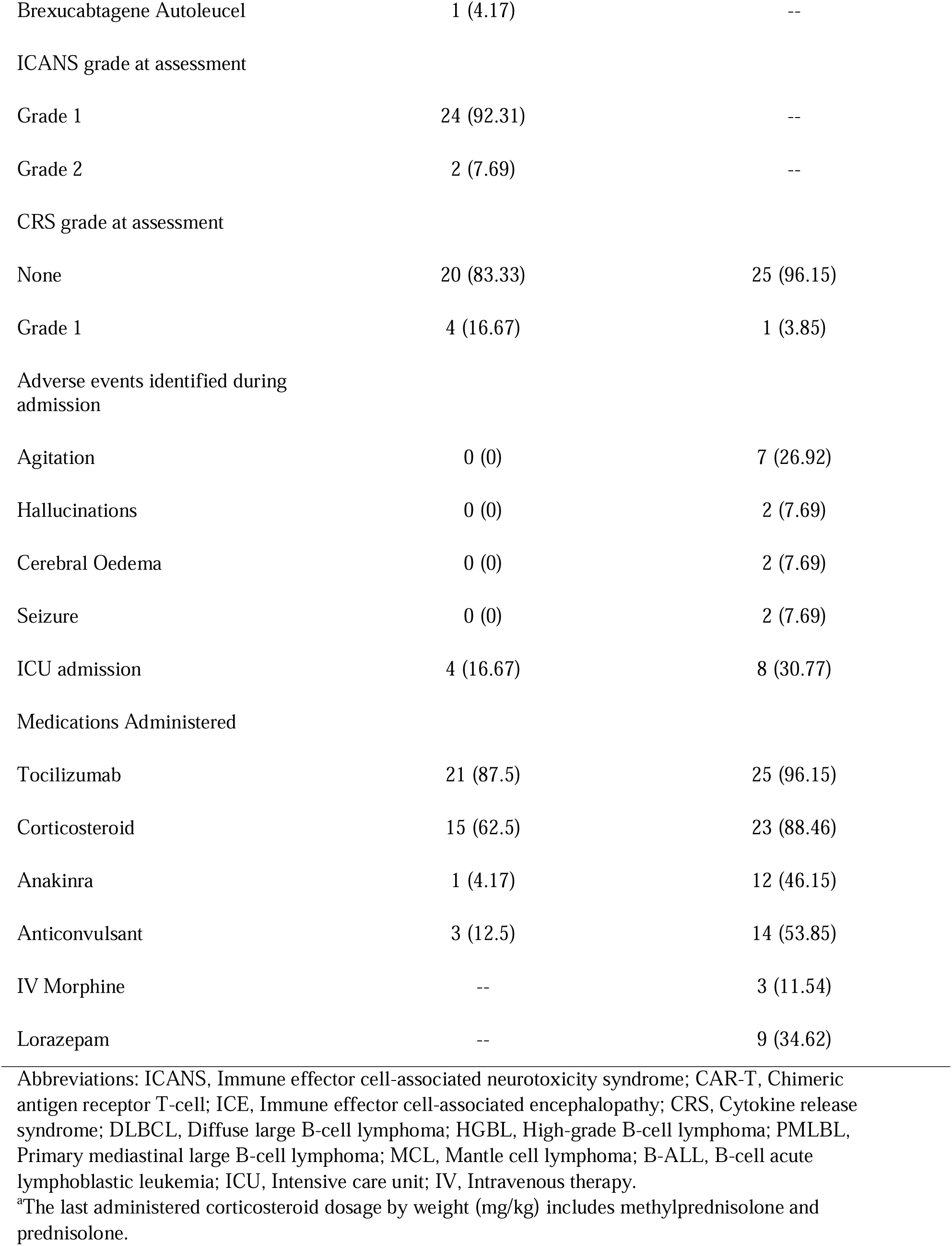
Clinicodemographic characteristics of CAR-T patients.

| Characteristic | Non-ICANS *M*(*SD*) | ICANS *M*(*SD*) |
| --- | --- | --- |
|  | *n* = 24 | *n* = 26 |
| Age (years) | 60.42 (13.86) | 62.08 (10.74) |
| Education (years) | 12.91 (2.86) | 12.15 (2.46) |
| Total length of inpatient stay (days) | 12 (4.14) | 17.92 (6.8) |
| Onset of ICANS (days) | -- | 5.21 (2.74) |
| Onset of CRS (days) | 3.9 (3.11) | 2.12 (1.59) |
| ICE score at the time of the neuropsychological examination | 9.96 (0.20) | 8.69 (0.74) |
| Last administered corticosteroid dosage by weight (mg/kg)<sup>a</sup> | 0.04 (0.06) | 0.07 (0.06) |
|  | N (%) | N (%) |
| Gender |  |  |
| Female | 10 (41.67) | 9 (34.62) |
| Male | 14 (58.33) | 17 (65.39) |
| Cancer Type |  |  |
| DLBCL | 20 (83.33) | 23 (88.46) |
| HGBL | 1 (4.17) | 2 (7.69) |
| PMLBL | 1 (4.17) | 1 (3.85) |
| MCL | 1 (4.17) | -- |
| B-ALL | 1 (4.17) | -- |
| CAR-T product |  |  |
| Axicabtagene Ciloleucel | 19 (79.17) | 25 (96.15) |
| Tisagenlecleucel | 4 (16.67) | 1 (3.85) |
| Brexucabtagene Autoleucel | 1 (4.17) | -- |
| ICANS grade at assessment |  |  |
| Grade 1 | 24 (92.31) | -- |
| Grade 2 | 2 (7.69) | -- |
| CRS grade at assessment |  |  |
| None | 20 (83.33) | 25 (96.15) |
| Grade 1 | 4 (16.67) | 1 (3.85) |
| Adverse events identified during admission |  |  |
| Agitation | 0 (0) | 7 (26.92) |
| Hallucinations | 0 (0) | 2 (7.69) |
| Cerebral Oedema | 0 (0) | 2 (7.69) |
| Seizure | 0 (0) | 2 (7.69) |
| ICU admission | 4 (16.67) | 8 (30.77) |
| Medications Administered |  |  |
| Tocilizumab | 21 (87.5) | 25 (96.15) |
| Corticosteroid | 15 (62.5) | 23 (88.46) |
| Anakinra | 1 (4.17) | 12 (46.15) |
| Anticonvulsant | 3 (12.5) | 14 (53.85) |
| IV Morphine | -- | 3 (11.54) |
| Lorazepam | -- | 9 (34.62) |

Abbreviations: ICANS, Immune effector cell-associated neurotoxicity syndrome; CAR-T, Chimeric antigen receptor T-cell; ICE, Immune effector cell-associated encephalopathy; CRS, Cytokine release syndrome; DLBCL, Diffuse large B-cell lymphoma; HGBL, High-grade B-cell lymphoma; PMLBL, Primary mediastinal large B-cell lymphoma; MCL, Mantle cell lymphoma; B-ALL, B-cell acute lymphoblastic leukemia; ICU, Intensive care unit; IV, Intravenous therapy.
<sup>a</sup>The last administered corticosteroid dosage by weight (mg/kg) includes methylprednisolone and prednisolone.

As shown in Figure 1, six items from the NUCOG meet the inclusion criteria for the computing of the cognitive screening index. These items were B5-Calculation, C1b-Verbal recall, C2-Spatial recall, D1-Sequencing, D2-Categorical fluency, and D3-Abstraction. The cognitive screening index was computed as the sum of the six items for each patient (*M* = 22.16, *SD* = 6.78, range = 4.5 – 33). The reliability of the cognitive screening index was high (GLB = .86).

**Figure 1.**
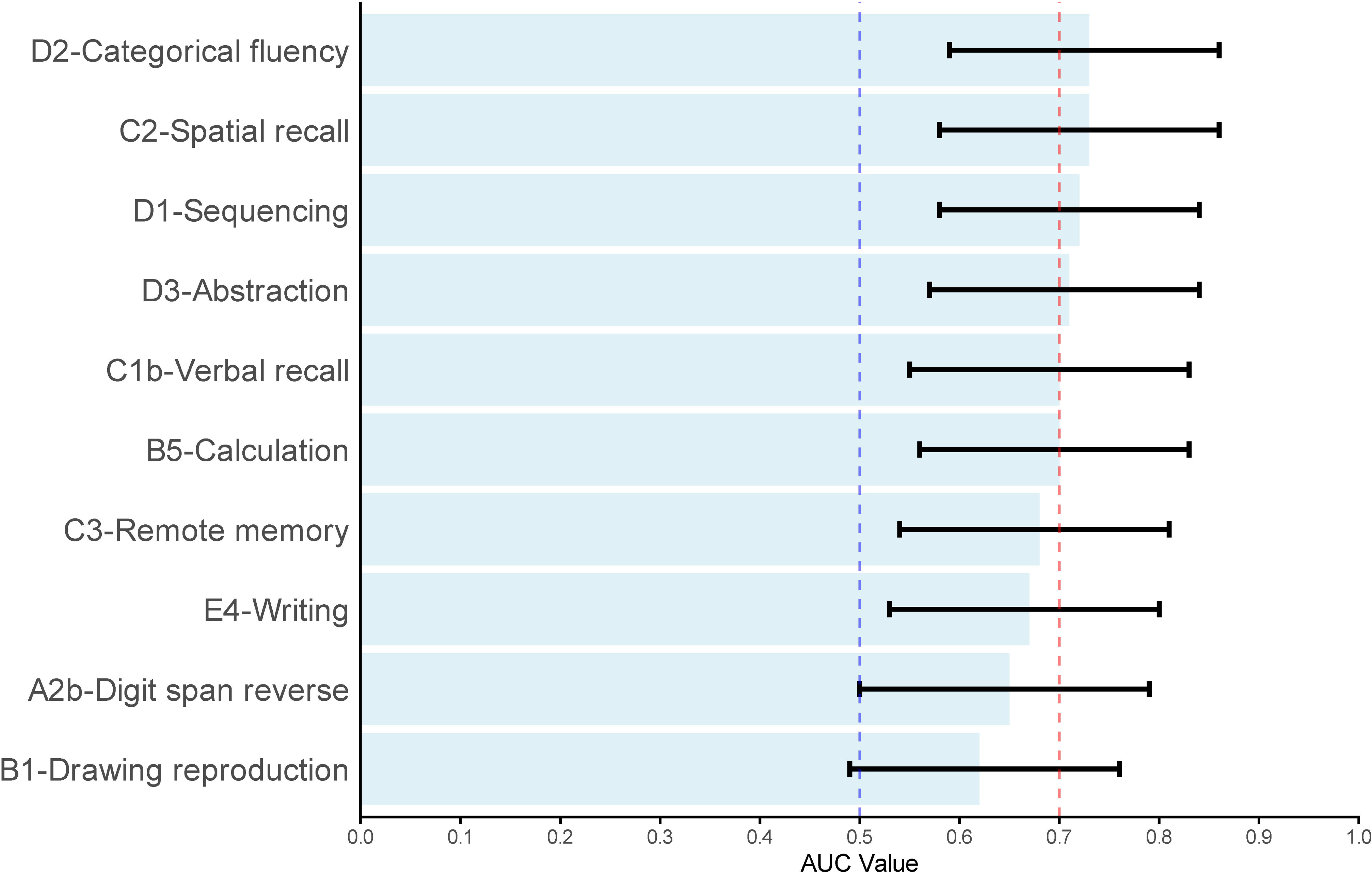
Area under the curve with bootstrapped confidence intervals of NUCOG Items. *Note*. NUCOG, Neuropsychiatry Unit Cognitive Assessment Tool; AUC, Area under the curve. The black line represents the AUC value of each test score with 95% bootstrapped confidence intervals. The blue dashed line represents the AUC value of .50. The red dashed line represents the threshold of “Acceptable” AUC value of .70. The figure depicts the top 10 NUCOG items with the highest AUC values.

The mean of the cognitive screening index was significantly lower in the ICANS group (*M =* 18.79, *SD* = 6.92) compared to non-ICANS group (*M =* 25.81, *SD* = 4.38), *t*(42.69) = 4.32, *p* < .001, *g* = 1.19. ROC curve analysis demonstrated that the cognitive screening index was able to discriminate between ICANS and non-ICANS cases (AUC = .81, 95% CIs [0.68, 0.92]). At an optimal threshold of 24, the cognitive screening index achieved sensitivity of .81, specificity of .71, a PPV of .75, and an NPV of .77. The computed second index is formulated based on the number of abnormal items as determined by Youden’s *J* statistic, where patients scored below the abnormal threshold across four out of the six items demonstrated sensitivity of .81, specificity of .79, a PPV of .81, and an NPV of .79. The individual item threshold is outlined in supplementary materials.

As shown in Figure 2, five neuropsychological test scores met the inclusion criteria for the computing of the neuropsychological index. These tests were TMT-A, HVLT-R delay trial, RCFT & TCFT, SDMT-Oral, and HVLT-R total learning. The neuropsychological index was computed by calculating the mean of these five tests for each patient (*M =* -1.24, *SD* = 1.14, range = -4.01 – 0.90). The reliability for the neuropsychological index was high (GLB = .83).

**Figure 2.**
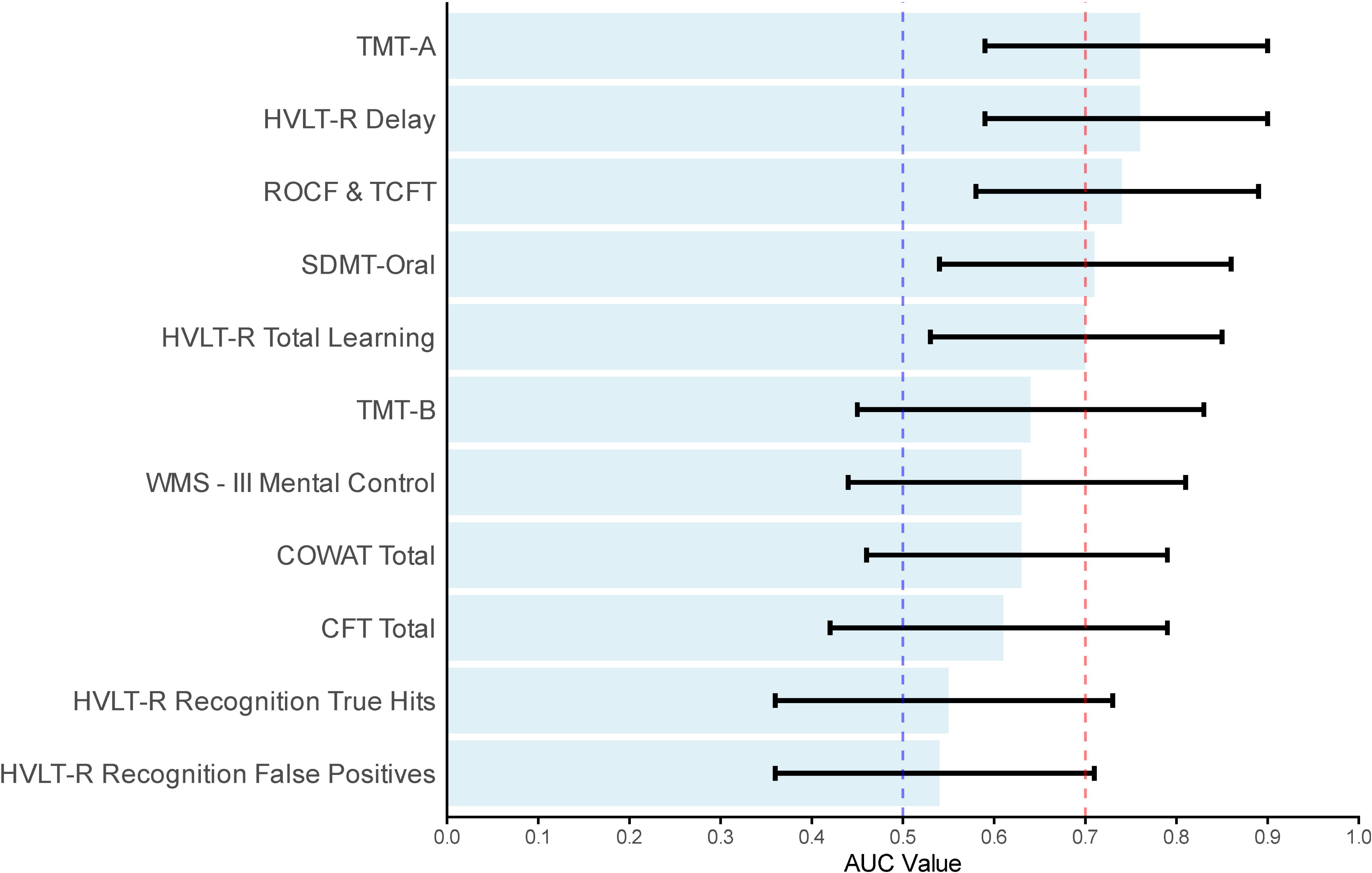
Area under the curve with bootstrapped confidence intervals of neuropsychological tests. *Note.* AUC = Area under the curve; TMT = Trails Making Test; HVLT-R = Hopkins Verbal Learning Testing - Revised; ROCF = Rey-Osterrieth Complex Figure; TCFT = Taylor Complex Figure Test; WMS-III = Weschler Memory Scale – III; SDMT-Oral = Symbol Digit Modalities Test – Oral; COWAT = Controlled Oral Word Association Test; CFT = Category Fluency Test. The black line represents the AUC value of test score with 95% bootstrapped confidence intervals. The blue dashed line represents the AUC value of .50. The red dashed line represents the threshold of the ‘Acceptable’ AUC value of .70.

The mean neuropsychological index was significantly lower in the ICANS group (*M =* -1.84, *SD* = 0.96) compared to the non-ICANS group (*M =* -0.61, *SD* = 0.97), *t*(44.88) = 4.36, *p* < .001, *g* = 1.25. ROC curve analysis demonstrated that the neuropsychological index was able to discriminate between ICANS and non-ICANS cases (AUC = .80, 95% CIs [0.57, 0.89]). At an optimal threshold of <= - 0.87, the neuropsychological index demonstrated sensitivity of .92, specificity of .62, a PPV of .73, and an NPV of .88. Using the -1.5 SD cut-off in line with clinical convention, the computed second index revealed that patients who scored below this cut-off on > 1 test demonstrated sensitivity of .81, specificity of .67, a PPV of .72, and an NPV of .76.

While the AUC value of the neuropsychological index was marginally higher (AUC = .81) than the cognitive screening index (AUC = .80), this difference did not reach statistical significance (*p* = .765). Figure 3A shows the ROC curve of the derived indices, and Figures 3B and 3C show the mean difference between the ICANS and the non-ICANS group on the derived indices.

**Figure 3.**
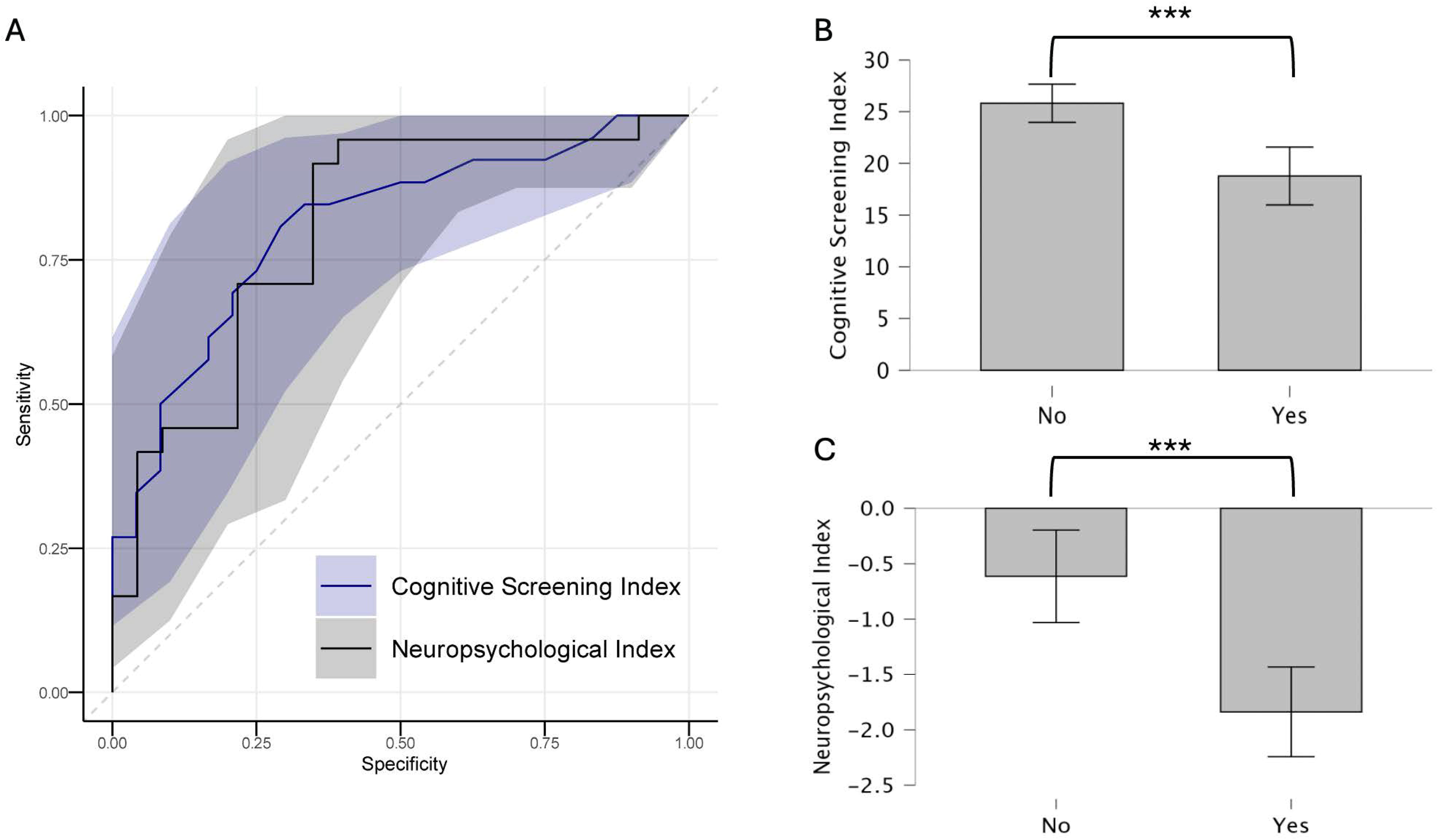
Neuropsychological and cognitive screening index in ICANS classification. *Note*. Figure 3A = ROC curves of the derived indices in ICANS classification. Figure 3B = Mean cognitive screening index scores of patients without and with ICANS. Figure 3C = Mean neuropsychological index scores of patients without and with ICANS. ICANS = immune effector cell-associated neurotoxicity syndrome. The error bars represent the 95% confidence intervals to the mean. ‘Yes’ = ICANS at assessment. ‘No’ = no-ICANS at assessment. \*\*\**p* < .001.

Hierarchical logistic regression was used to determine whether the neuropsychological index improved detection of ICANS over and beyond the cognitive screening index alone. For the baseline model, only the cognitive screening index was entered. This model was statically significant, χ^2^ (1) = 17.6, *p* < .001, Nagelkerke *R*^2^ = .42. As a second step, the neuropsychological index was added to the model (χ^2^ (2) = 22.4, *p* < .001, Nagelkerke *R*^2^ = .51), which significantly improved the model fit (χ^2^ (2) = 4.87, *p* < .01) and explained 9% more variance compared to the baseline model. In the full model, every one-score increase in the cognitive screening index decreased the odds of the CAR-T patient being classified as ICANS by 1.19 times. Every unit of z-score increase in the neuropsychological index decreased the odds of CAR-T patients being classified as ICANS by 2.57 times.

The cognitive screening index and the NUCOG total score (AUC = .79, 95% CIs [0.66, 0.91]) did not statistically differ in the ability to detect ICANS (*p =* .824). The ROC curves of the cognitive screening index and the NUCOG total score in classification of ICANS are provided in Figure S2 in the supplementary materials. Comparison of the cognitive screening index and the NUCOG total score in classification of controls to CAR-T patients is also outlined in supplementary materials.

As shown in Table 2, the derived indices significantly correlated with clinical characteristics, including the ICE score at assessment and the total length of inpatient stay. Higher scores on both indices were associated with a higher ICE score at assessment and a shorter total length of inpatient stay. Neither index correlated with the last administered corticosteroid dosage.

**Table 2.**
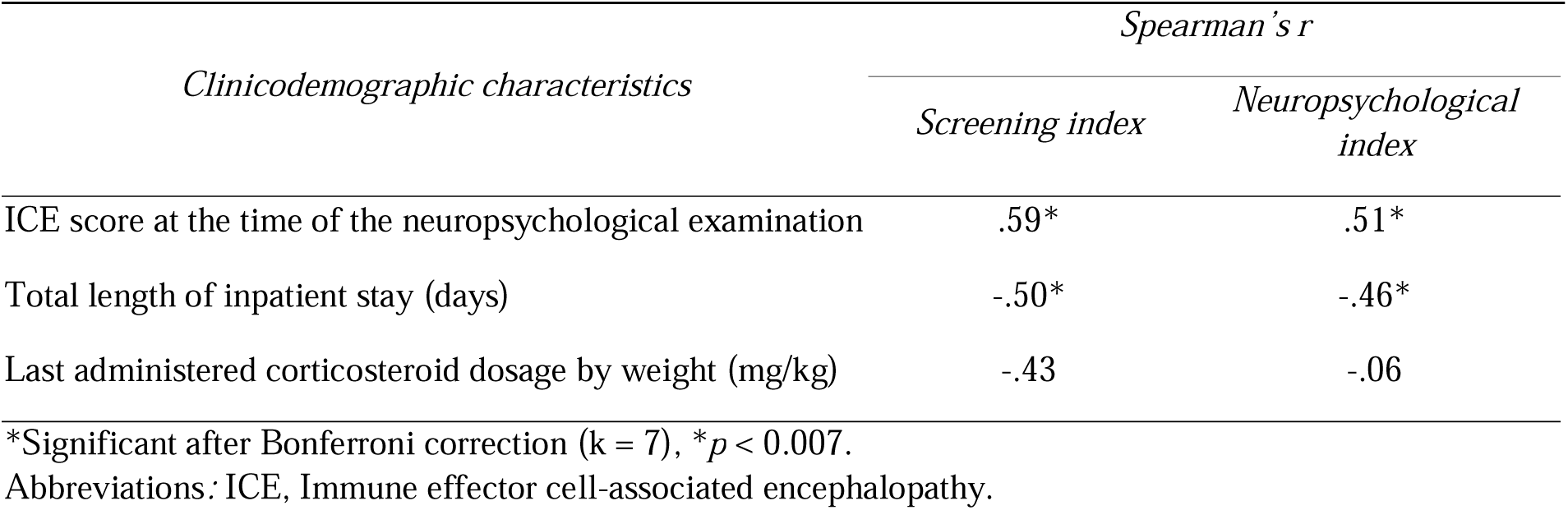
Correlations between key clinicodemographic characteristics and derived indices.

## Discussion

This study aimed to develop a novel screening instrument sensitive to ICANS-related cognitive dysfunction. As expected, ICANS patients were abnormal on a subset of screening items and neuropsychological tests compared to non-ICANS patients. The derived cognitive screening and neuropsychological indices demonstrated good psychometric reliability and classification accuracy of ICANS status. While both indices achieved similar classification accuracy, the neuropsychological index captured additional information over and beyond the cognitive screening index alone on ICANS status, confirming its incremental validity. Compared to the NUCOG total score, the cognitive screening index achieved a similar accuracy in identifying ICANS. Both indices correlated with the total length of inpatient stay and the ICE score at assessment. Neither index correlated with corticosteroid doses administered prior to the assessment.

The pattern of abnormal neuropsychological test scores in the ICANS group indicates impairment in cognitive domains of processing speed, attention, executive function, verbal memory, and visuoconstructional function. This pattern is consistent with acute cognitive symptoms of ICANS reported in existing studies, which include attentional, executive, and memory dysfunction.^8–10^ In the present study, ICANS patients exhibited abnormal performance on all tests of processing speed, attention, and executive function with or without psychomotor components. Processing speed is an important component of the attentional system.^28^ The slower processing speed and attention impairments observed in the study suggests that ICANS as a cognitive syndrome may be primarily driven by slowed speed and attentional dysfunction.

The present findings show that the cognitive screening index presents as a viable approach to detection of cognitive changes in ICANS. Early detection of ICANS is crucial for timely intervention that could improve clinical outcomes.^29^ The limitations of the currently used ICE score in accurately detecting early cognitive signs of ICANS have been highlighted by Herr et al^11^ and Yuen et al.^10^ As shown in Figure 4, the present study combined the most useful cognitive screening items into a new instrument, the ICE-COG, which examines aspects of cognition different from those captured by the ICE score. Thus, it is hypothesised that the ICE-COG could improve early detection of ICANS, which needs to be validated in future research using another clinical cohort.

**Figure 4.**
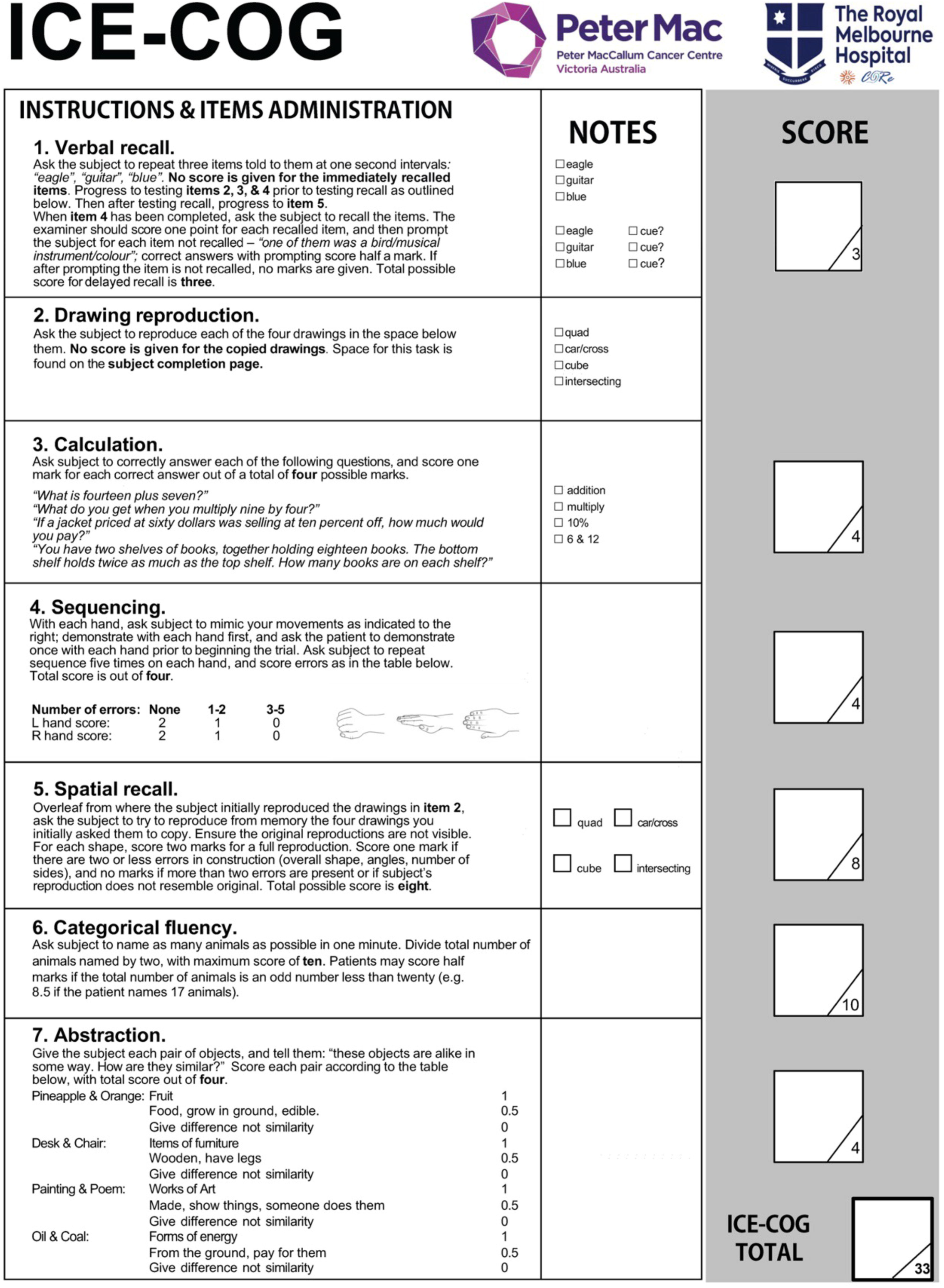
Immune effector cell-associated encephalopathy cognitive assessment tool (ICE – COG)

The neuropsychological index showed superiority in identifying ICANS patients over the cognitive screening index alone. This suggests that neuropsychological examination remains the benchmark for diagnosing acute cognitive change. Neuropsychological assessment might not be available for all CAR-T patients when hospital resources are limited. Combining the approaches highlighted in the current study may offer an optimal pathway for acute monitoring of CAR-T patients. The ICE-COG could be routinely administered to screen for cognitive change acutely after CAR-T. Patients with abnormal scores would then be referred for a neuropsychological assessment for cognitive characterisation.

The derived indices were associated with a longer length of inpatient stay and a lower ICE score at assessment. A lower ICE indicates more severe cognitive complications,^3^ and a prolonged inpatient stay is often a result of adverse events like ICANS.^30^ The direction of these associations was consistent with expectation, thereby establishing the criterion validity of the indices.

There were several limitations in the present study. Patients were classified as ICANS or non-ICANS based on the ASTCT consensus grading criteria, which included the ICE score.^3^ Classification accuracy and psychometric properties of the derived indices were computed based on this classification. This study was the first to explore a new approach to monitoring cognitive change after CAR-T. Future studies, however, are needed to validate the indices in an independent cohort. Based on the psychometric properties, it is hypothesised that the ICE-COG could aid in early detection of ICANS, provide information beyond the ICE score, and direct earlier intervention. Future research should compare the ICE score with the ICE-COG to examine which instrument would be the first to identify cognitive change. Future research may also investigate whether administering the ICE-COG immediately post-infusion in patients with normal ICE scores could predict the onset and severity of ICANS.

The present study did not examine different grades of ICANS severity due to a limited sample size. Patients with severe ICANS may also present with neurological complications, such as cerebral oedema and seizures, and therefore would likely present with acute cognitive characteristics different to those observed in cases of mild ICANS.^31^ Future studies should compare acute cognitive presentation across severity grades. Corticosteroid dosage was included to control for any effects of corticosteroid treatment, which is routinely used to manage ICANS.^32,33^ The lack of an association suggests that the use of corticosteroids was not a meaningful confounder and that higher doses were not associated with greater cognitive impairment. Future studies should, however, reexamine an association between acute corticosteroid dosage and cognitive presentation in a larger cohort.

## Conclusion

The current study aimed to develop a novel screening instrument sensitive to ICANS-related cognitive dysfunction. Our findings showed that ICANS patients were disproportionally impaired in processing speed, attention, and executive function compared to non-ICANS patients. The derived ICE-COG demonstrated good clinical utility in identifying cognitive impairments in ICANS. The instrument can be used by the treating team to monitor CAR-T patients acutely after infusion. Patients who screen abnormally on the ICE-COG can subsequently be referred to neuropsychology for further assessment. Future studies are needed to validate the instrument in other cohorts and examine whether it is sensitive to early cognitive changes in ICANS. Capturing early cognitive symptoms in these patients will allow for early intervention, which could improve clinical outcomes.

## Data Availability

Anonymised data can be shared on reasonable request from qualified investigators.

## Acknowledgements

We acknowledge and thank CAR-T patients and their families for their participation. We also thank medical and administrative teams at Peter MacCallum Cancer Centre and Royal Melbourne Hospital for their patient care and logistical contribution to this study.

