## Supplementary Materials for "Brief assessment of cognition in immune effector cell-associated neurotoxicity syndrome"

**Figure S1**

*Flowchart of inpatient neuropsychological examination*

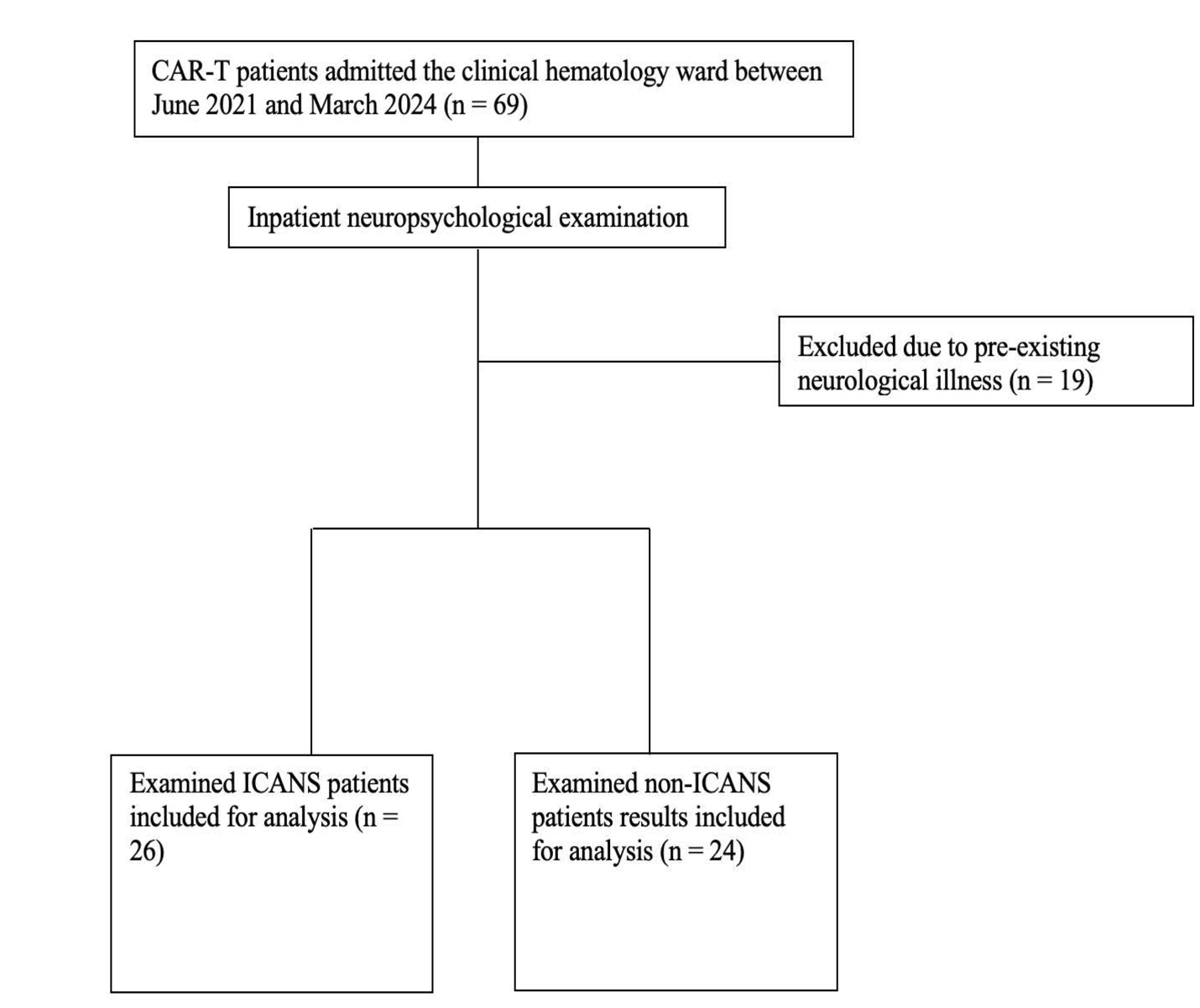
*Note.* CAR-T = Chimeric antigen receptor T-cell; ICANS = Immune effector cell-associated neurotoxicity syndrome.

**Figure S2**

*Cognitive screening index and NUCOG total score in classification of ICANS status*

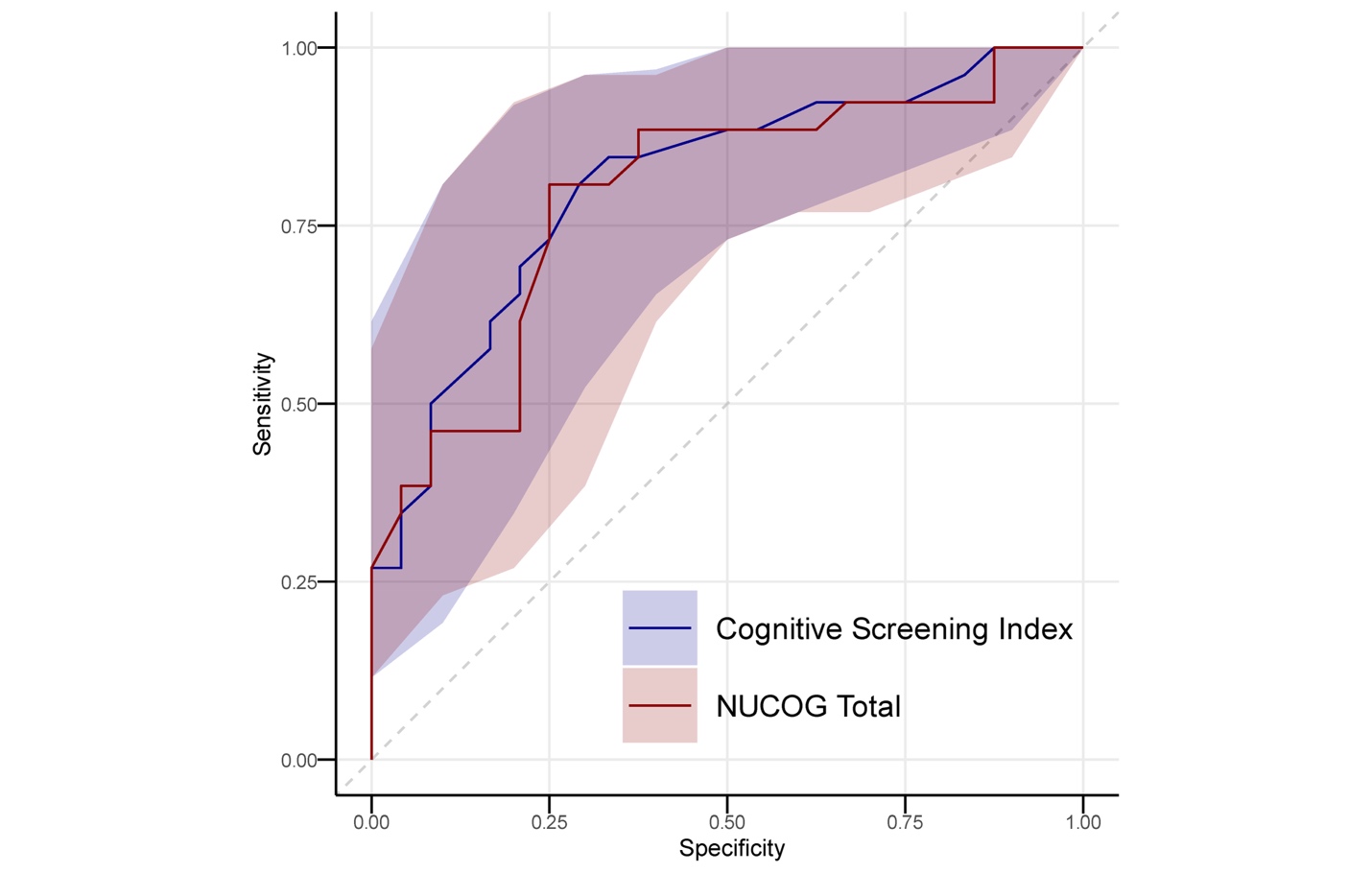
*Note*. ICANS = Immune effector cell-associated neurotoxicity syndrome; NUCOG = Neuropsychiatry Unit Cognitive Assessment Tool. The shaded region depicts the bootstrapped confidence intervals of the respective measure.

**Figure S3**

*Cognitive screening index and NUCOG total score in classification of CAR-T patients and controls*

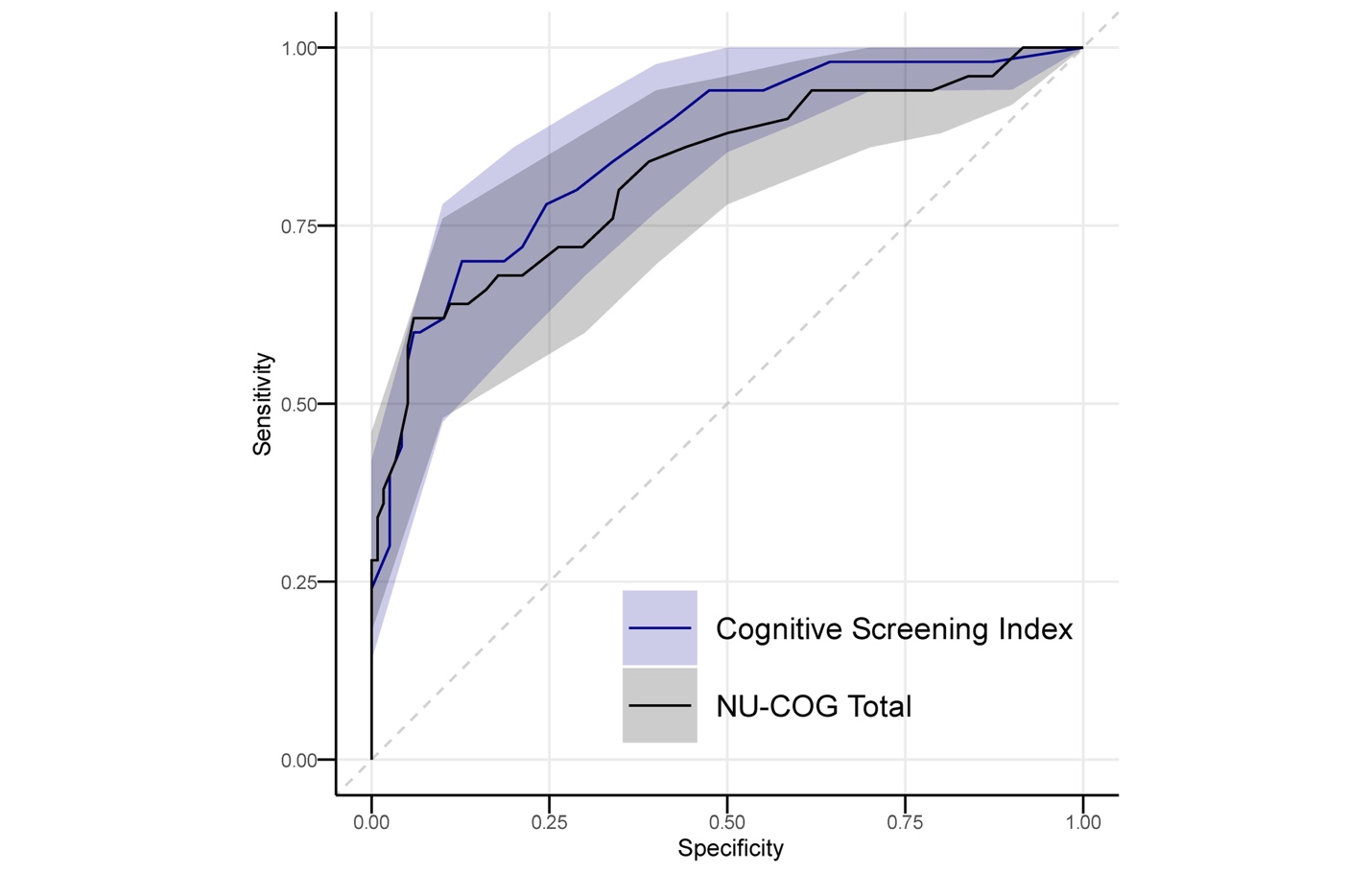
*Note*. CAR-T = Chimeric antigen receptor T-cell; NUCOG = Neuropsychiatry Unit Cognitive Assessment Tool. The shaded region depicts the bootstrapped confidence intervals of the respective measure.

† The AUC value of the cognitive screening index was marginally higher than the NUCOG total in classification of CAR-T patients to healthy controls but did not reach statistical significance (*p* = .08).

In the classification of ICANS patients and healthy controls, the cognitive screening index demonstrated (AUC = .94, 95% CIs [0.89 – 0.98]) a sensitivity of .85, a specificity of .94, a PPV of .76, and an NPV of .97 at the threshold of 24. In classification of non-ICANS patients to hospitalised controls, the cognitive screening index (AUC = .77, 95% CIs [0.66 – 0.86]) demonstrated a sensitivity of .75, a specificity of .66, a PPV of .31, and an NPV of .93 at the threshold of 29. Consistent with the slopes of the ROC curves (Figure S4), permutation testing revealed that the ROC curve of ICANS patients to hospitalised controls was superior in classification accuracy compared to the ROC curve of non-ICANS patients to hospitalised controls (*p* < *.*01).

**Figure S4**

*ROC curves of the cognitive screening index in classification of ICANS patients and non-ICANS patients to healthy controls*

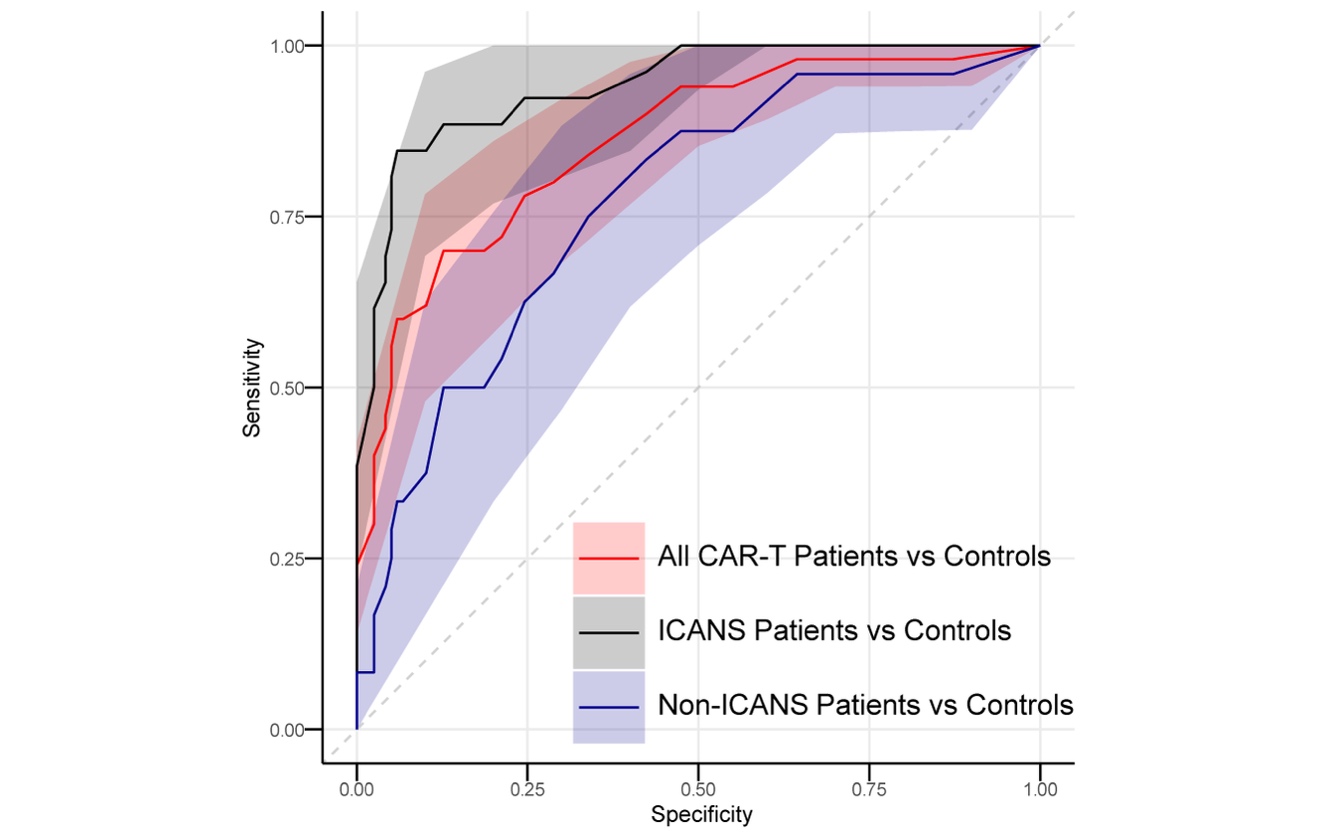

*Note*. ROC = Receiver operator characteristic; CAR-T = Chimeric antigen receptor T-cell; ICANS = Immune effector cell-associated neurotoxicity syndrome. The shaded region depicts the bootstrapped confidence intervals of the respective measure.

**Table S5.**

*Normative Data and Test Description*

| Instrument | Normative Data |
| --- | --- |
| Weschler Memory Scale III-Mental Control (WMS-III) | Weschler D. *Wechsler Memory Scale–Third Edition (WMS-III).* The Psychological Corporation; 1997. |
| Symbol Digital Modalities Test-Oral (SDMT-Oral) | Smith, A. *Symbol digit modalities test (SDMT) manual (revised)* Western Psychological Services; 1982. |
| Hopkins Verbal Learning Test–Revised (HVLT-R) | Benedict RHB, Schretlen D, Groninger L, Brandt J. Hopkins Verbal Learning Test – Revised: Normative Data and Analysis of Inter-Form and Test-Retest Reliability. *Clin Neuropsychol*. 1998;12(1):43-55. doi: 10.1076/clin.12.1.43.1726 |
| Controlled Oral Word Association Test (COWAT) & Category Fluency Test (CFT). | Tombaugh TN, Kozak J, Rees L. COWA by education level in adults. In: Spreen O, Strauss E, eds. *A Compendium of Neuropsychological Tests: Administration, Norms, and Commentary.* Oxford University Press; 1998:453. |
| Trail Making Test A & B (TMT) | Tombaugh T. Trail Making Test A and B: Normative data stratified by age and education. *Arch Clin Neuropsychol*. 2004;19(2):203-214. doi: 10.1016/s0887-6177(03)00039-8 |
| Rey-Osterrieth Complex Figure and Taylor Complex Figure Test (ROCF & TCFT) | Mitrushina M, Boone KB, Razani J, D'Elia LF. *Handbook of normative data for neuropsychological assessment*. Oxford University Press; 2005;783. |

*Note*. TMT = Trails Making Test; HVLT-R = Hopkins Verbal Learning Testing - Revised; ROCF = Rey-Osterrieth Complex Figure; TCFT = Taylor Complex Figure Test; WMS-III = Weschler Memory Scale – III; SDMT-Oral = Symbol Digit Modalities Test – Oral; COWAT = Controlled Oral Word Association Test; CFT = Category Fluency Test.
